# Transcriptomic Signaling Pathways involved in a naturalistic model of Inflammation-related Depression and its remission

**DOI:** 10.1101/2020.09.21.20196592

**Authors:** Marie-Pierre Moisan, Aline Foury, Sandra Dexpert, Steve W Cole, Cédric Beau, Damien Forestier, Patrick Ledaguenel, Eric Magne, Lucile Capuron

## Abstract

This study aimed at identifying molecular biomarkers of inflammation-related depression in order to improve diagnosis and treatment. We performed whole-genome expression profiling from peripheral blood in a naturalistic model of inflammation-associated major depressive disorder (MDD) represented by comorbid depression in obese patients. We took advantage of the marked reduction of depressive symptoms and inflammation following bariatric surgery to test the robustness of the identified biomarkers. Depression was assessed during a clinical interview using Mini-International Neuropsychiatric Interview and the 10-item, clinician administered, Montgomery-Asberg Depression Rating Scale. From a cohort of 100 massively obese patients we selected 33 of them for transcriptomic analysis. Twenty-four of them were again analyzed 4-12 months after bariatric surgery. We conducted differential gene expression analyses before and after surgery in unmedicated MDD and non-depressed obese subjects. We found that TP53 (Tumor Protein 53), GR (Glucocorticoid Receptor) and NF*κ*B (Nuclear Factor kappa B) pathways were the most discriminating pathways associated with inflammation-related MDD. These signaling pathways were processed in composite z-scores of gene expression that were used as biomarkers in regression analyses. Results showed that these transcriptomic biomarkers highly predicted depressive symptom intensity at baseline and their remission after bariatric surgery. While inflammation was present in all patients, GR signaling overactivation was found only in depressed ones where it may further increase inflammatory and apoptosis pathways. In conclusion, using an original model of inflammation-related depression and its remission without antidepressants, we provide molecular predictors of inflammation-related MDD and new insights in the molecular pathways involved.

## Introduction

Over the past two decades, compelling evidence has emerged to highlight the role of inflammation in the onset and perpetuation of major depressive disorders (MDD). Epidemiological and clinical studies have repeatedly shown the high prevalence of depressive symptoms in chronic inflammatory disorders. Furthermore, chronic treatment with the pro-inflammatory cytokine, interferon-alpha, in medically ill patients was found to be responsible for the development of major depression in over 30% of patients [1, 2]. Consistent with these data, a recent meta-analysis confirmed that MDD is associated with elevated levels of pro-inflammatory cytokines, such as interleukin (IL)-6 and tumor necrosis factor (TNF)-α [3]. Additionally, genetic studies have revealed the role of immune genetic variants in MDD [4]. Interestingly, inflammation has been recently hypothesized to also play a role in antidepressant treatment resistance, as patients presenting signs of systemic low-grade inflammation display a poor response to classical antidepressants, such as selective serotonin reuptake inhibitors [5]. Thus, a better understanding of the molecular mechanisms underlying inflammation-related depression is needed. In particular, the identification of biomarkers specific to this depression subtype is required for improving its diagnosis and treatment.

Transcriptomic analysis from peripheral blood has successfully been used to reveal biological pathways linked to MDD [6,7]. A recent study examined the implication of specific molecular pathways in inflammation-related depression by measuring self-reported depressive symptoms 2h after endotoxin injection in healthy subjects [8]. However, hypothesis-free transcriptomic analyses have not been performed specifically in diagnosed inflammation-associated MDD patients. To address this issue, we used a naturalistic model of inflammation-related depression represented by comorbid depression in obese patients. Indeed, obesity is a chronic medical condition characterized by a low-grade inflammatory state associated with an increased prevalence of depression [9]. Remarkably, systemic inflammation, reflected by increased levels of C-reactive protein levels, predicts obesity-related depressive symptoms better than metabolic health per se [10]. Interestingly, bariatric surgery-induced weight loss was found to correlate with a reduction of both inflammation and depressive symptoms [11, 12].

In this study, we sought to identify transcriptional control pathways associated with inflammation-related depression in unmedicated obese patients. For this purpose, gene expression profiling of peripheral blood was performed and compared between depressed and non-depressed severely obese patients before and after bariatric surgery, with the hypothesis that these biological pathways would be changed after surgery-induced weight loss and subsequent decrease in inflammation. Upstream regulator analyses were employed to uncover the molecular signaling pathways that differentiate depressed from non-depressed subgroups of patients. Finally, the most discriminating signaling pathways identified were processed in composite z-scores of gene expression in order to be used as biomarkers predicting depressive symptoms.

## Methods and Materials

### Patients

One hundred severely or morbidly obese patients (body mass index (BMI) > 35 kg/m2) awaiting bariatric surgery were recruited from the services of digestive and bariatric surgery at two private clinics (Tivoli and Jean Villar clinics, Bordeaux, France). Patients were scheduled to receive either a sleeve gastrectomy or a gastric bypass. Patients were chosen on the basis of their levels of depressive symptoms in order to cover a range from non-depressed to clinically depressed patients. Seventy-three percent of patients (n=24) were followed up at 4-12 months after surgery (mean time at follow-up 6.5 months). Exclusion criteria were: age > 65 years old; acute or chronic inflammatory conditions (other than obesity); current treatment with antidepressants or any other psychotropic drug; current diagnosis of psychiatric disease (except for major depression); and/or severe medical illness. The study was approved by the local Committee for the Protection of Persons (Bordeaux, France). All patients provided written informed consent after reading a complete description of the study.

### Clinical evaluation

DSM criteria for current major depression were determined using the Mini-International Neuropsychiatric Interview (MINI) administered during a semi-structured interview by trained raters at baseline and after bariatric surgery [13]. Concomitantly, the intensity of depressive symptoms was assessed using the 10-item, clinician administered, Montgomery-Asberg Depression Rating Scale (MADRS) [14].

### Genome-wide transcriptional profiling

Blood samples were collected twice, before (n=33) and after surgery (n=24) in study participants. Five ml of venous blood was collected per patient on vacuum tubes (PAXgene blood RNA system; PreAnalytiX GmbH, Hombrechtikon, Switzerland). The samples were maintained at room temperature for > 2 h as required for stabilization of RNA and then kept at −20°C until RNA extraction. Total RNA was extracted using the PAXgene Blood RNA Kit (Qiagen, Courtaboeuf, France) according to the manufacturer’s protocol. Quality of the total RNA was assessed using RNA Nano chips on a Bioanalyser 2100 (Agilent, Boeblingen, Germany). All samples had an RNA Integrity Number (RIN) score > 8.0. RNA concentrations were measured with a Nanodrop spectrophotometer. For each sample, Cyanine-3 (Cy3) labeled cRNA was prepared from 200 ng of total RNA using the One-Color Quick Amp Labeling kit (Agilent) according to the manufacturer’s instructions, followed by RNA clean-up using Agencourt RNAClean XP (Agencourt Bioscience Corporation, Beverly, Massachusetts). Dye incorporation and cRNA yield were checked using Dropsense™ 96 UV/VIS droplet reader (Trinean, Belgium). Six hundred ng of Cy3-labelled cRNA were hybridized on the microarray slides following the manufacturer’s instructions. Immediately after washing, the slides were scanned on Agilent G2505C Microarray Scanner using Agilent Scan Control A.8.5.1 software and fluorescence signal extracted using Agilent Feature Extraction software v10.10.1.1 with default parameters. Gene expression profiles of the 57 samples (33 before and 24 after surgery) were analyzed in a single batch at the GeT-TRiX facility (GenoToul, Génopole Toulouse, France) using Agilent SurePrint G3 Human GE V2 8×60K (Design 039494) microarrays following the manufacturer’s instructions. Microarray data and experimental details are available in NCBI’s Gene Expression Omnibus [15] and are accessible through GEO Series accession number GSE99725.

### sPLS regression

Sparse Partial Least Square (sPLS) regression [16] using the R package, mixOmics (http://www.mixOmics.org) was performed to select the expressed genes that correlated the best to MDD diagnosis or MADRS items. From the 2000 probes found highly correlated with a correlation coefficient r>0.5, 1474 unique genes were identified.

### Differential Gene Expression analyses

Raw data (median of pixel intensity) were analyzed using R (www.r-project.org, R v. 3.1.2) Bioconductor packages (www.bioconductor.org, v 3.0,([17]) as described in GEO entry GSE99725. Briefly, raw data were filtered, log2 transformed, corrected for bath effects (washing and labeling serials) and normalized using the quantile method [18]. A first exploratory and statistical analysis showed a possible correlational structure among gene expression, which could negatively impact the multiple testing procedures. We applied the FAMT method [19] to reduce the dependence structure. A model was fitted using the limma lmFit function [20] considering “patients” as blocking factor for the time pairs using duplicate correlation function. A correction for multiple testing was then applied using the Benjamini-Hochberg procedure [21] for false discovery rate (FDR). Probes with FDR ≤ 0.05 were considered differentially expressed between conditions.

### Impact of blood cell proportions in gene expression profiles

We assessed whether the observed gene expression changes in the different analyses were related to changes in cell proportions in the blood samples using the Cell-type Computational Differential Estimation CellCODE R package [22].

### Transcriptional pathways identification, comparison analysis and composite z-scores analyses

The biological pathways involved in MDD were predicted from lists of differentially expressed genes using the “Upstream Regulator Analysis” from Ingenuity Pathway Analysis (IPA, Qiagen, USA).

To calculate the composite z-scores representing a given signaling pathway, we first extracted the list of target genes for a given transcription factor from ChIP-seq experiments available in ChEA 2016, an integrative database of public ChIP-seq data (available in http://amp.pharm.mssm.edu/Enrichr/). Then, from the list of target genes for a given transcription factor, we selected the ones that were expressed in our dataset. For each of the patients, the expression data for each gene was first standardized by calculating a z-score,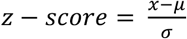 where *x* is the gene expression data, the mean of the gene expression for the population (all patients of the study) and *σ* the standard deviation for the population. For each patient, the composite z-score for a given transcription factor was then calculated as the mean of the target genes’ z-scores.

ANOVA and Multivariate regression analyses were done with the software Statistica and the graphs designed with GraphPadPrism. A p value of p<0.05 was considered as significant.

## Results

### Characteristics of participants

The general characteristics of the participants are described in Table 1. Since study participants were chosen at baseline to cover a range from non-depressed to clinically depressed subjects, the percentage of patients with MDD in the present study (42.42%) is higher than what is generally found in the obese population. There were no differences in terms of age (36.57±11.22 vs 39.95±11.87, t=0.83, p=0.41), gender (85.71% females MDD, 94.74% females non MDD, Fischer’s exact test p=0.56) and BMI (41.64±1.17 vs 42.33±5.70, t=0.38, p=0.71) between non-depressed *versus* depressed patients. As expected, BMI as well as depressive symptoms were strongly reduced after surgery in the 24 patients with available data at follow up (BMI: paired t test t=24.65 p<0.0001; MADRS scores: paired t test t=5.26, p<0.0001). Similarly, the percentage of patients with MDD was significantly decreased after surgery compared to baseline (from 42.42% to 0%, Fisher exact test p<0.0006).

**Table 1:** Characteristics of study participants

|  | Baseline n=33 | Post-surgery n=24 |
| --- | --- | --- |
| Age, mean [SD] | 38.52 ± 11.54 | - |
| Women, n [%] | 30 [90.9%] | 21 [87,5%] |
| BMI, kg/m<sup>2</sup> | 42.09 ± 5.10 | 31.27 ± 5.10 |
| MDD, n [%] | 14 [42.42%] | 0 |
| MADRS score | 12.21±7.28 | 4.13 ± 4.68 |

### Gene expression analyses

As a first exploration of the peripheral blood gene expression data from the 33 obese patients at baseline, we performed a sPLS regression analysis with the variables related to depressive symptoms. The heatmap (Fig.1A) illustrates the strength of correlation between the 2000 most correlated probes and the variables related to depressive symptomatology. Strong correlations with gene expression were found with total MADRS score and MDD diagnosis. Not surprisingly, correlation was the weakest with the MADRS item measuring reduced appetite. When the selected variable was total MADRS score, the 2000 most correlated probes corresponded to 1474 unique genes, 428 down-regulated and 1046 up-regulated. When this gene dataset was submitted to IPA, the top canonical pathway identified was the Glucocorticoid Receptor (GR) signaling pathway (p value 2.13 10^−5^ with an overlap of 12.5%), while the top upstream regulators were cystatin D (CST5), tumor protein 53 (TP53) and hepatocyte nuclear factor4 (HNF4A) (p= 1.09 10^−8^, p=9.8 10^−8^ and p= 6.10 10^−7^ respectively).

**Figure 1:**
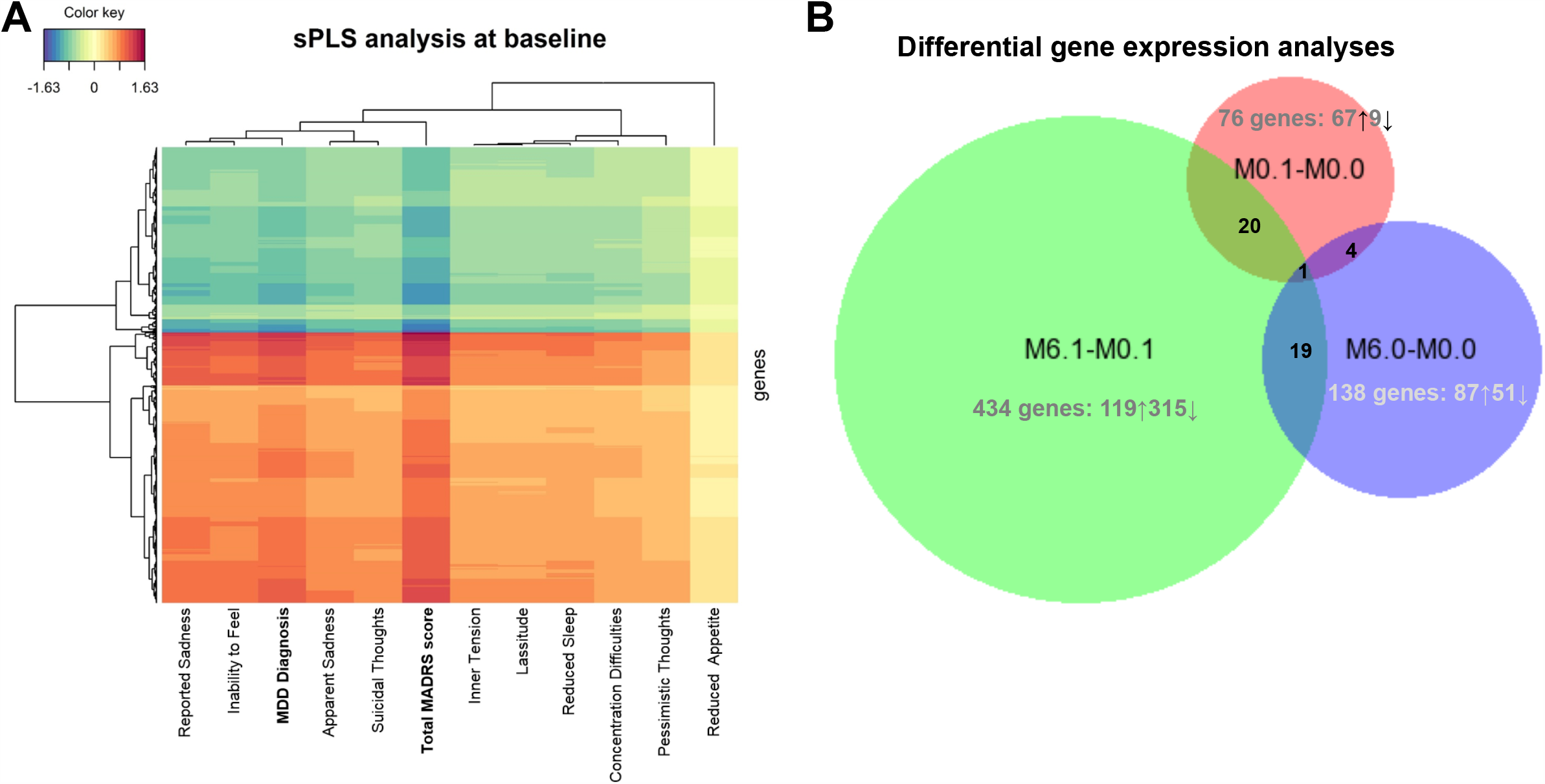
Gene expression analyses at baseline and after bariatric surgery in MDD and non-depressed patients.

Then, we performed 4 differential gene expression analyses where we compared the gene expression of MDD to non-depressed patients at baseline (M0.1-M0.0), MDD patients after *versus* before bariatric surgery (M6.1-M0.1), non-depressed patients after *versus* before surgery (M6.0-M0.0) and lastly, MDD *versus* non-depressed (as they were at baseline) after surgery (M6.1-M6.0).

At a false discovery rate of 5%, the three first analyzes provided a number of differentially expressed genes (DEG) as illustrated in Fig.1B, with Venn diagram proportional to the number of DEG. For the later analysis (M6.1-M6.0), no significant DEGs were detected as expected since patients were no longer depressed in either group. We found no significant changes in immune cell subtypes in the different analyses using CellCODE (Table S1). The list of DEG for each analysis, and the corresponding fold change, is presented in Table S2.

### Upstream regulator analyses

To identify the transcriptional pathways associated with MDD in these patients, we perform upstream regulator analyses from each list of DEG using IPA. The lists of upstream regulators for each analysis are provided in Table S3. At a cutoff p value of overlap p<0.001, only one upstream regulator, TP53, was significant for the (M0.1-M0.0) analysis, with 17 downstream target genes. For the (M6.1-M0.1) analysis, 67 upstream regulators were detected, the three most significant were respectively dehydrotestosterone (p= 4.25 ⨯ 10^−7^, an androgen receptor agonist), TP53 (p= 1.6 ⨯ 10^−6^) and dexamethasone, a specific glucocorticoid receptor agonist (p= 2.84 ⨯ 10^−6^,). TP53 and dexamethasone showed the highest number of downstream target genes with 67 and 71 target genes respectively. Interestingly, a mechanistic network was revealed that linked dexamethasone and TP53 along with other upstream regulators of this dataset (Fig.2A). The NF*κ*B (Nuclear Factor kappa B) complex appears as an important hub between dexamethasone and TP53 in this network. For the (M6.0-M0.0) analysis, 7 upstream regulators were found significant but all of them with a low number of target genes, GATA1 (GATA binding protein 1) having the highest number with 9 target genes. To identify the upstream regulators that were in common or different between MDD and non-depressed patients and explaining the changes after surgery, a comparison analysis was performed (Fig.2B). This analysis highlighted that among the 10 regulators sorted by activation z-score, several cytokines pathways (IL-4, IL15, IL2, IL-1β) were down-regulated in both MDD and non-depressed patients. Remarkably, dexamethasone was found down-regulated in the (M6.1-M0.1) analysis only.

**Figure 2:**
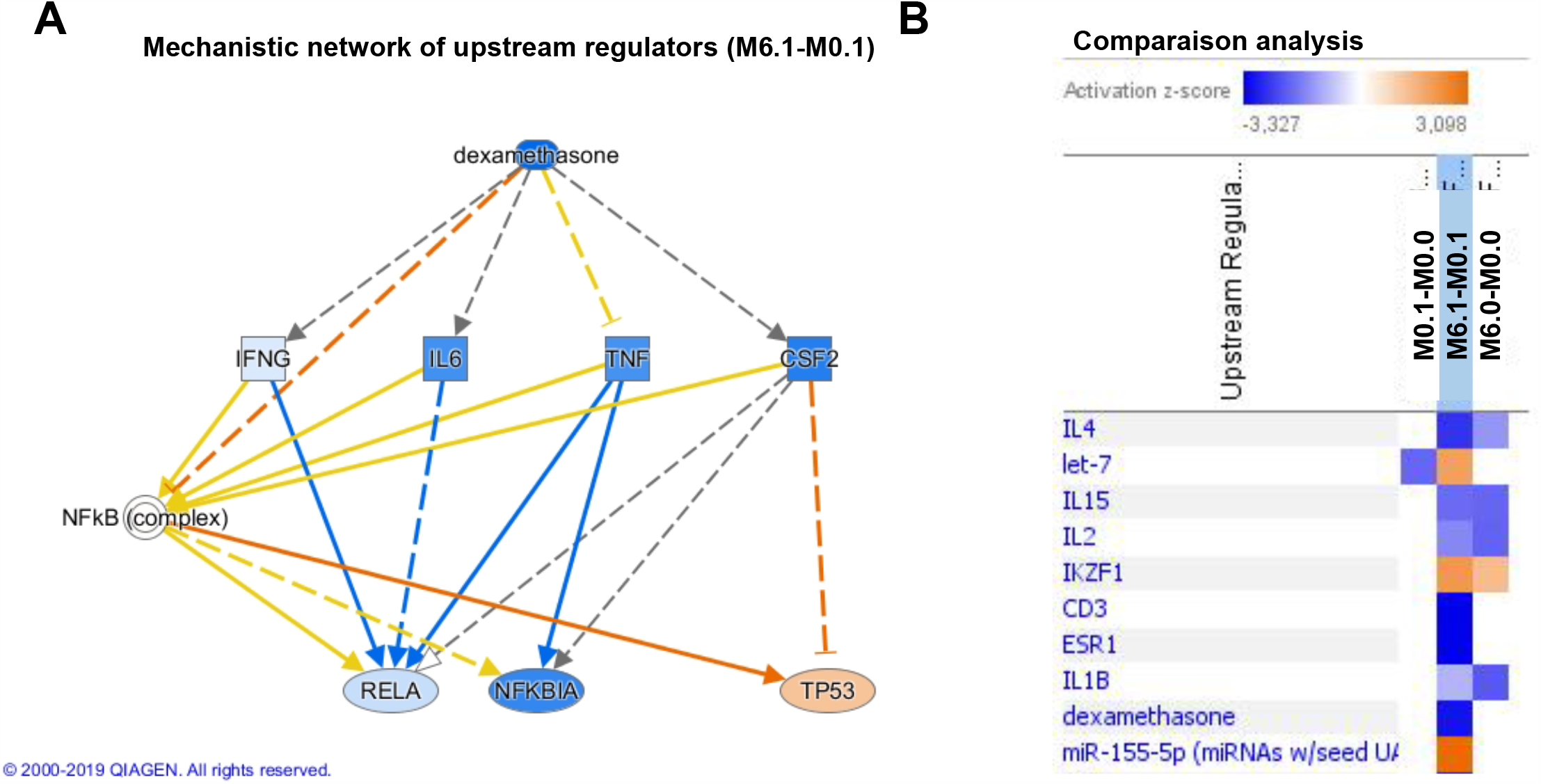
Upstream regulator analyses using Ingenuity Pathway Analysis.

### Transcriptomic pathway biomarkers of inflammation-related depression

The upstream regulator analyses above, in accordance with the sPLS regression, pinpointed dexamethasone/GR, NF*κ*B and TP53 as the most important and convergent transcriptional regulators associated with MDD. To assess whether these pathways could serve as biomarkers predicting inflammation-related depression in our sample, we calculated for each of these regulators a composite z-score of gene expression using all their target genes, identified from ChIP-seq experiments and expressed on the microarrays (see Methods for details). The lists of target genes representing NR3C1 (for dexamethasone/GR), RELA (for NF*κ*B) and TP53 pathways and used to calculate the composite z-scores are given in Table S4. Results from a 2-way ANOVA indicated that MDD patients could be differentiated from non-depressed patients at baseline by the composite z-scores of TP53, NR3C1 and RELA, which were normalized after surgery when patients were no longer depressed (Figure 3) (for each graph 2-way ANOVA interaction (MDD ⨯ Surgery) p<0.01, Bonferroni post hoc tests: MDD vs. non-depressed patients before surgery p<0.001, MDD vs. non-depressed patients after surgery p=ns, MDD patients before surgery vs. after surgery p<0.001).

**Figure 3:**
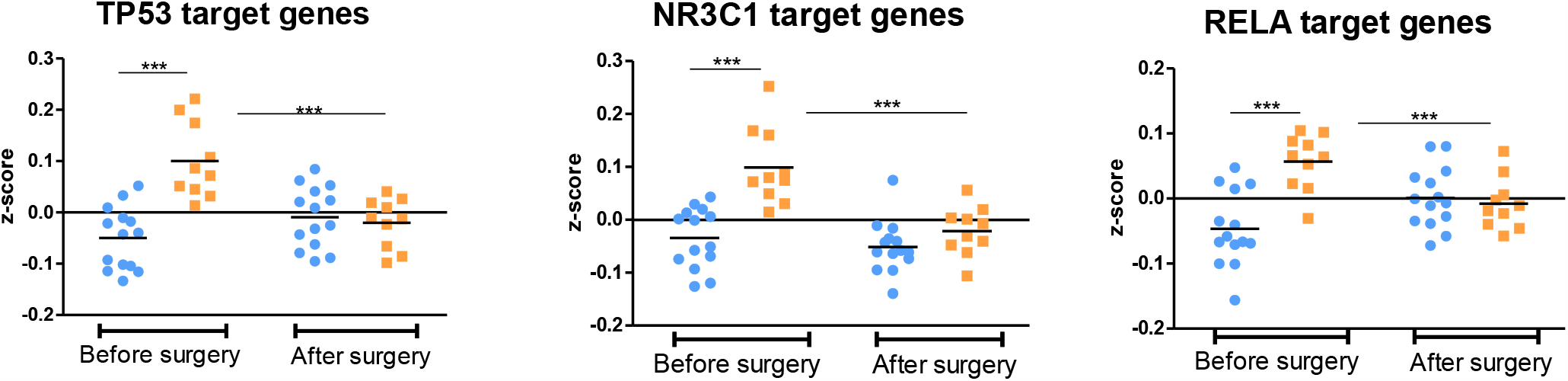
TP53, NR3C1 and RELA pathways differentiate MDD (orange symbols) from non-depressed patients (blue symbols) at baseline and are normalized after surgery. 2-way ANOVA followed by Bonferroni post hoc tests, ***p<0.001.

Furthermore, we tested by linear regression analysis whether these composite z-scores could predict the intensity of depressive symptoms at baseline using total MADRS scores (Fig.4A). Indeed, highly significant linear relationships were detected for each of the 3 pathways. TP53 showed the strongest relationship (β=0.80 F_(1,31)_=56.8 p<10-7), followed by NR3C1 (β=0.79 F_(1,31)_=50.3 p<10-7) and RELA (β=0.78.5 F_(1,31)_=49.8 p<10-7). These associations remained significant after adjusting for gender, age, and BMI (Table S5). Using the same composite z-scores in regression analyses with BMI, we found no relationship (Figure S1) suggesting the specificity of the transcriptomic biomarkers for depressive symptoms. Since depressive symptoms were considerably reduced in most obese patients after surgery, we hypothesized that the difference in composite z-scores before and after surgery should predict the decrease in MADRS scores from baseline to post-surgery. Consistent with this hypothesis, multivariate linear regression analyses showed significant linear relationship for the 3 transcription factor composite z-scores (β=0.65 F_(1,22)_=16.4 p<0.001 for TP53, β=0.47 F_(1,22)_=6.34 p<0.02 for NR3C1 and β=0.49 F_(1,22)_=7.19 p<0.015 for RELA) (Fig.4B).

**Figure 4:**
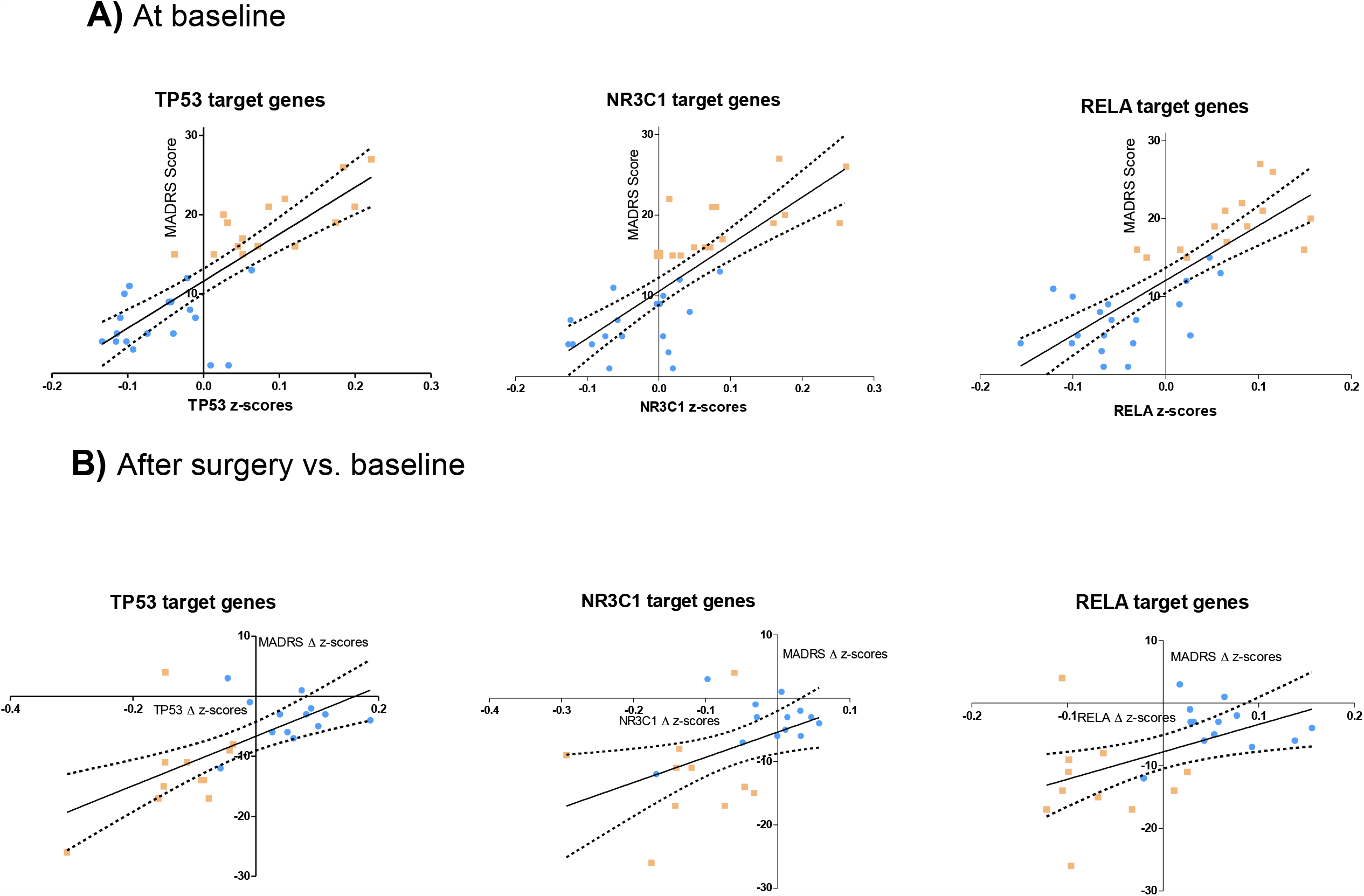
TP53, NR3C1 and RELA pathways predict the intensity of inflammation-related depressive symptoms at baseline (A) and their decrease after bariactric surgery (B). Multivariate linear regression analyses. MDD patients (orange symbols), non MDD patients (blue symbols).

## Discussion

Low-grade-inflammation in obesity originates primarily from the adipose tissue, in which immune cells accumulate and secrete inflammatory factors [9, 23]. The resulting systemic inflammation is thought to contribute to increased neuroinflammatory processes that subsequently drive behavioral alterations, including depressive symptoms [11]. In the present study, we used this model of inflammation-related depression to reveal the transcriptomic pathways involved, taking advantage of the marked reduction of depressive symptoms and inflammation following bariatric surgery. Our transcriptomic and bioinformatic analyses identify TP53, NR3C1 and NF*κ*B as the most important upstream regulators predicting depression in obese patients. Indeed, NR3C1/Glucocorticoid Receptor and TP53 signaling pathways were revealed as linked to depressive symptoms by a sPLS analysis using the 33 patients at baseline and the involvement of these 2 upregulators were confirmed in the differential gene expression analysis comparing post-surgery *versus* baseline in MDD patients. Regarding NFKB, it was first identified as an important intermediary in a mechanistic network including NR3C1 and TP53.

TP53 is known as a tumor suppressor, pro-apoptotic protein which levels are altered in response to cellular stress. Increased apoptotic stress was reported previously in animal models of stress-induced depression and in MDD patients [24, 25]. For example, a transcriptomic analysis of postmortem prefrontal cortex tissues from patients with a history of MDD revealed altered expression of apoptosis factors together with increased cytokines expression [26]. The importance of TP53 pathway itself was described in an animal model of trauma by transcriptional analysis from blood, amygdala and hippocampus [27] and in a genetic study where polymorphisms within TP53 gene were found associated with MDD in a Slovak population [28]. Interestingly, TP53 was also shown to be increased in the adipose tissue of obese mice and responsible for cytokine production and senescence-like changes thereby contributing to insulin resistance [29]. Our finding that TP53 pathway is involved in inflammation-related depression in obese individuals is consistent with these data. It is tempting to postulate that TP53 induces the secretion of pro-inflammatory factors in adipose tissue, which in turn triggers neuroinflammation and subsequent behavioral alterations.

Since MDD patients were not different from non-depressed patients in terms of BMI, it is very likely that other factors besides adiposity explain increased TP53 and NF*κ*B signaling in MDD patients. Our analysis put forward NR3C1/GR as one possible factor since it came out in both sPLS and differential gene expression analyses as an important upstream regulator associated with MDD. Furthermore, in the comparison analysis, dexamethasone appeared as a significant upstream regulator in obese MDD patients but not in obese non-depressed patients. Evidence from the literature strongly suggests a role for GR in the development of MDD and its related neurobiological disturbances. Indeed, MDD is often preceded by a history of stress and chronic elevated glucocorticoid levels, which contribute to reduced hippocampal volume and impaired hippocampal neurogenesis commonly found in MDD [30]. Recently, GR signaling was found as the most significant pathway associated with trauma-related individual differences in a preclinical model as revealed by gene expression analysis. It was also the most significant transcription factor convergent across blood and brain tissues. Furthermore, a translational study discovered that GR sensitivity had a crucial role in antidepressant treatment response in MDD patients and in a mouse model of depression [31]. As for inflammation-related depression, a recent study examined candidate transcriptomic pathways contributing to depressed mood in healthy subjects treated acutely with endotoxin. NF*κ*B and CREB [cAMP Responsive Element Binding Protein] signaling were found increased in subjects who developed depression but GR signaling was decreased [8]. This study confirmed earlier reports showing for example that activation of inflammatory processes in MDD is associated with a reduced GR*α/β* expression ratio in monocytes [32,33] or that dexamethasone-stimulated gene expression in peripheral blood is lower in depressed patients than in healthy subjects [34]. Although these data may seem contradictory to ours for GR, it could be explained by the anti and pro-inflammatory actions of glucocorticoids which depend on the timing of exposure [35,36,37]. Along this line, we can hypothesize that obese patients with depressive symptoms may have been exposed to more stress and elevated glucocorticoid levels before developing obesity, which would result in increased inflammation during fat accumulation. In support of this hypothesis, we showed in a previous study that patients who developed MDD during chronic interferon-α treatment displayed, as soon as the first administration of the cytokine (at the time where they were not depressed yet), an exaggerated ACTH and cortisol responses, suggestive of a sensitized stress response system [38].

Interaction between GR and TP53 has been reported in neuronal cell cultures where dexamethasone through GR enhances TP53 activity by increasing its nuclear translocation and transcriptional activity [39]. Induction of TP53 by dexamethasone was also found in cultured tenocytes and in tendon biopsies from patients treated with dexamethasone [40] supporting the role of GR in TP53 induction in vivo.

A number of studies have utilized gene expression profiles to investigate the determinants of MDD. Although few associated genes are found in common across studies, all of them implicate immune pathways, such as NF*κ*B or pro-inflammatory cytokines [41-46], oxidative stress or apoptosis [26,43,47] signaling pathways. These convergent findings confirm that transcriptomic pathways represent stronger biomarkers than individual genes, probably due to statistical issues, effects of cellular heterogeneity and temporal dynamics, as well as multifactorial gene regulation, as discussed before for stress genomics [48], and heterogeneity of depression types.

In most studies, the transcriptomic pathways are identified from the differentially expressed genes by bioinformatic prediction based on transcription binding motif using various software (e.g., DAVID[49], TELiS [50]] or literature curation (IPA)). Here, we used a new approach relying on ChIP-seq experiments. By this method, we believe that we captured a closer reflection of the transcriptional pathways involved. Indeed, the composite z-score of gene expression representing the pathways of TP53, GR and RELA were highly predictive of MADRS scores in our patient sample. Furthermore, the same composite z-scores were also predictive of the remission of depressive symptoms after bariatric surgery, supporting the validity of these biomarkers.

In conclusion, this study highlights the importance of TP53, NF*κ*B and GR signaling pathways in a clinical model of inflammation-related MDD and their potential as predictive biomarkers to identify individuals vulnerable to this type of depression. While replication of these findings in larger cohorts and in non-obese patients with inflammation-associated depression is needed, the present data provide insights of the molecular mechanisms involved in inflammation-associated depression that can be used to improve diagnosis and treatment.

## Data Availability

all the data are in supplemental material

## Funding and Disclosures

This work was supported by funds from the French National Research Agency, ANR (ANR-11-JSV1-0006, LC), and the *Fondation pour la Recherche Médicale* (DPP20151033943, LC).

The authors declare no conflict of interest.

## Acknowledgements

The authors thank Claire Naylies and Yannick Lippi for their contribution to microarray fingerprints acquisition and microarray data analysis carried out at GeT-TRiX Genopole Toulouse Midi-Pyrénées facility, Dr. J.J. Peroua, Dr. M. Salzmann-Lovato and the medical assistants for their help with patient recruitment, and the patients for their participation in the study.

## Authors contribution

MPM & LC design the study, AF, SD, CB, DF, PL & EM acquired the data, MPM, AF & LC analyzed and interpreted the data, MPM, SWC & LC wrote the manuscript. All authors approved the final version of the manuscript.

Supplementary Information accompanies this paper at (https://doi.org/………)

## Notes

### Competing Interest Statement

The authors have declared no competing interest.

### Author Declarations

The study was approved by the local Committee for the Protection of Persons (Bordeaux,France)

